# Epidemiology of sleep disorders during COVID-19 pandemic: A systematic scoping review protocol

**DOI:** 10.1101/2020.05.17.20104794

**Authors:** Samia Tasnim, Mariya Rahman, Priyanka Pawar, Liye Zou, Abida Sultana, E. Lisako J. McKyer, Ping Ma, Md Mahbub Hossain

## Abstract

**Background:** The coronavirus disease (COVID-19) is impacting human health globally. In addition to physical health problems, a growing burden of mental health problems has become a global concern amid this pandemic. Sleep disorders are major mental health problems associated with increased psychosocial stressors; however, no research synthesis is available on the epidemiology of sleep disorders. In this systematic scoping review, we aim to assess the current evidence on the epidemiological burden, associated factors, and interventions from the existing literature on sleep disorders.

**Methods:** We will search seven major health databases and additional sources to identify, evaluate, and synthesize empirical studies on the prevalence and correlates of sleep disorders and available interventions addressing the same. We will use the Joanna Briggs Institute Methodology for Scoping Review and report the findings using the Preferred Reporting Items for Systematic Reviews and Meta-Analyses extension for Scoping Reviews (PRISMA-ScR) checklist.

**Conclusion:** This review will identify the epidemiological burden of and interventions for sleep disorders. The findings of this review will be widely communicated with the research and professional community to facilitate future research and practice.

## Introduction

The novel coronavirus disease (COVID-19) is critically affecting not only the physical health but also mental health globally (Hossain et al., 2020a; Rajkumar, 2020; Sultana et al., 2020). Many studies have reported a growing burden of a wide range of mental disorders including sleep disorders (Rajkumar, 2020; Zhang et al., 2020). Psychosocial stressors like anxiety, stress, changed lifestyle with limited social support, and fear may affect the pattern of sleep among individual often leading to sleep disorders (Krystal, 2012; Staner, 2003). As there is a growing concern on mental health problems during COVID-19 (Hossain et al., 2020b; Rajkumar, 2020), research synthesis may play a critical role in understanding the burden of those problems and addressing the same. To the best of our knowledge, there is a scarcity of evidence on the magnitude of sleep disorders among individuals affected by this pandemic. This scoping review aims to address this knowledge gap through systematically evaluating the current evidence on the epidemiological burden of sleep disorders, associated factors, and interventions addressing the same.

## Review Questions

1. What is the epidemiological burden of sleep disorders in different populations during COVID-19?
2. What are the factors associated with sleep disorders during COVID-19?
3. What are the available interventions for addressing sleep disorders amid COVID-19?

## Inclusion criteria

### Participants

In this scoping review, we include participants irrespective of their sociodemographic conditions. This makes our review inclusive for all types of participants who fulfill remaining criteria of this review.

### Concepts

This review will focus on sleep disorders, which can be defined by the International Classification of Diseases or Diagnostic and Statistical Manual of Mental Disorders (American Psychiatric Association, 2013; World Health Organization, 2010). Moreover, sleep abnormalities expressed as insomnia, excessive sleepiness, poor sleep quality, and abnormal events that occur during sleep will also be considered as sleep disorders in this review (Thorpy, 2012). Studies reporting the prevalence, incidence, frequency, score, level or any forms of quantitative assessment of sleep-related conditions will be included in this review.

### Context

This review especially emphasizes on COVID-19 as the context. Therefore, studies conducted among populations affected by COVID-19 (doctors or patients) or population at risk (general population who could have had infected with COVID-19) will be considered in this review. Moreover, studies without mentioning relevance to COVID-19 would be excluded from this review.

### Types of sources

This review will include original studies, cross-sectional or longitudinal in nature, published as peer-reviewed journal articles. Studies published in English language will be included in this review. Therefore, unpublished works, non-original articles (for example, letters with no original research reports, editorials, reviews, commentaries etc.), non-peer reviewed articles, and studies in languages other than English will be excluded from this review.

## Methods

This scoping review with be conducted using the Joanna Briggs Institute (JBI) methodology for scoping reviews (Peters et al., 2015). Moreover, the findings of this review will be reported using the Preferred Reporting Items for Systematic Reviews and Meta-Analyses extension for Scoping Reviews (PRISMA-ScR) checklist (Tricco et al., 2018). The protocol of this review has been registered with The Open Science Framework (Tasnim et al., 2020).

### Search strategy

We will search MEDLINE, Embase, PubMed, Academic Search Ultimate, Cumulative Index to Nursing and Allied Health Literature (CINAHL), Web of Science, and APA PsycInfo databases using the keywords with Boolean operators as mentioned in Table 1.

**Table 1:** Search terms for this scoping review.

| **Search terms** | **Keywords (searched within titles, abstracts, and subject headings fields)** |
| --- | --- |
| **1** | “COVID-19” OR “2019-nCoV” OR “2019 coronavirus” OR “Wuhan coronavirus” OR “Wuhan virus” OR “Wuhan pneumonia” OR “2019 novel coronavirus” OR “novel coronavirus” OR “SARS-CoV-2” |
| **2** | “sleep” OR “sleep disturbances” OR “sleep disorders” OR “sleep problems” OR “insomnia” OR “circadian rhythm” OR “restless leg syndrome” OR “sleep apnea” OR “narcolepsy” OR “daytime dysfunction” OR “daytime sleepiness” |
| **3** | prevalence OR incidence OR epidemiology OR frequency OR case OR rate OR occurrence OR correlates OR determinants OR predictors OR "risk factors" OR "associate factors" OR interventions OR treatment OR therapy OR management |
| **Final search strategy** | 1 AND 2 AND 3 |

The keywords will be searched within the titles, abstracts, and subject headings fields. Moreover, as COVID-19 literature started to get published since late 2019, we limit our search within 2019 and 2020. Moreover, we will search the reference lists and citing articles in Google Scholar to identify additional articles that may meet our criteria.

### Study selection

After searching the databases, we will import all the citations to Rayyan QCRI, a cloud-based software for systematic reviews. Two authors (ST and MR) will independently evaluate those citations using the inclusion criteria of this review as stated earlier. At the end of independent screening, potential conflicts will be reviewed and resolved based on discussion in the presence of a third author (MMH). Finally included citations will undergo full-text assessment and articles meeting all the criteria will be considered for data extraction. A flow chart of the study selection process is depicted in Figure 1.

**Figure 1:**
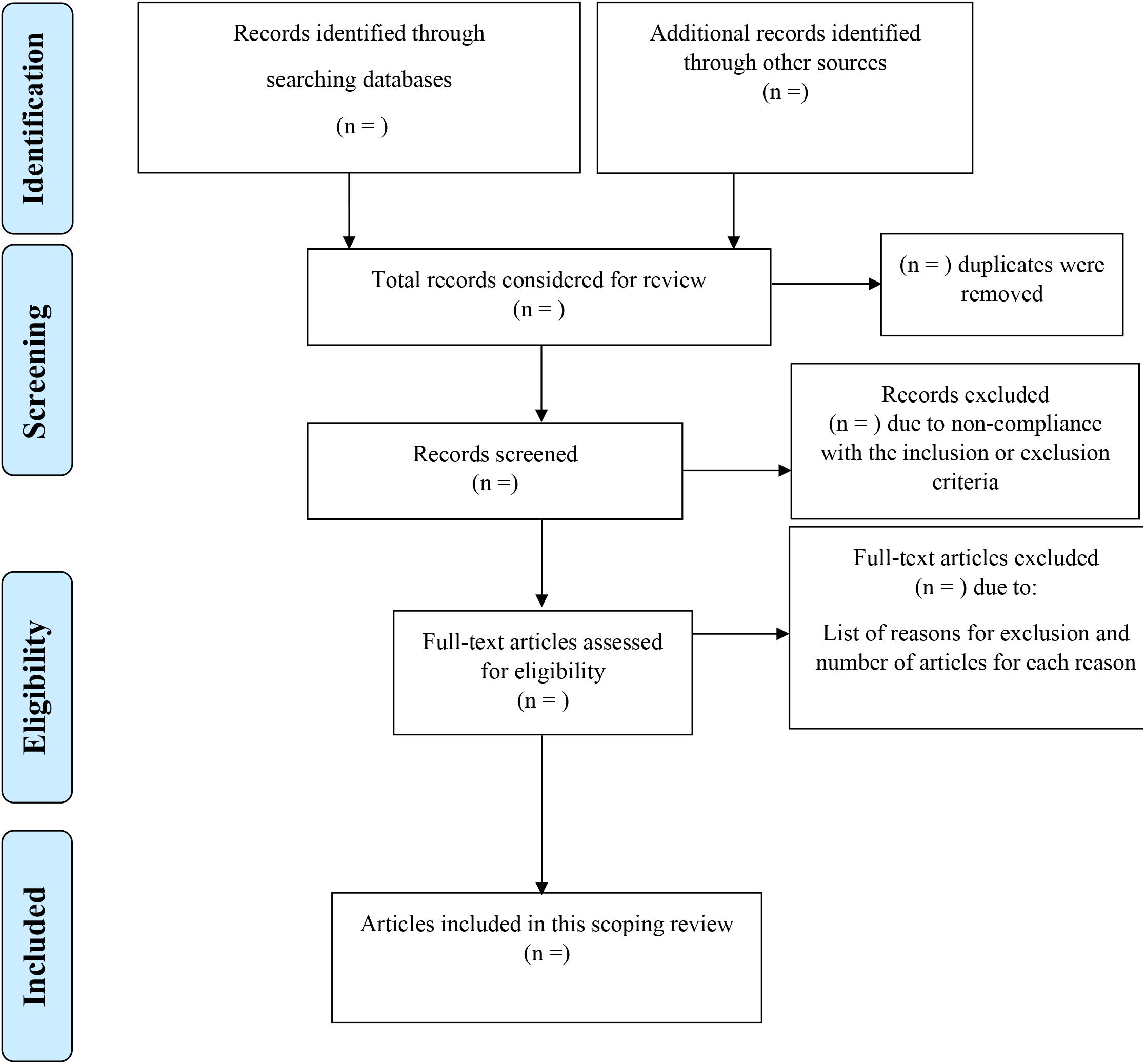
Flow diagram of the systematic scoping review.

### Data extraction

A data extraction form is prepared as shown in Table 2. Two authors will independently extract data using this checklist. At the end of this phase, data for each article will be reviewed by a third author to check consistency and potential conflicts will be re-assessed by three reviewers and a consensus will be made based on discussion.

**Table 2:** Data extraction instrument.

| **Study information** | **Title** |
| --- | --- |
|  | Author(s) |
|  | Year of publication |
|  | Country of origin |
|  | Type of the study |
|  | Objective of the study |
| **Information on study participants** | Sample size |
|  | Sample characteristics |
|  | Recruitment strategy |
| **Epidemiological data** | Methodology of the study |
|  | Timeframe of the study (with duration of exposure or intervention, if available) |
|  | Prevalence or quantitative burden of sleep disorders |
|  | Factors (if reported) associated with sleep disorders |
|  | Interventions (if reported) addressing sleep disorders (components and outcomes, if available) |
| **Citation of the article** | Full citation of the article including digital object identifier (DOI) |
| **References identified from the cited articles** | Articles that were cited in an article or forward citations of that article that may meet the criteria of this review |

### Data presentation

Data extracted from the included articles will be narratively synthesized and presented using tables and a commentary on key findings on the study characteristics, samples, and epidemiological findings as the quantitative burden and associated factors of sleep disorders during COVID-19 and interventions addressing the same. As per the JBI methodology, scoping reviews do not aim to evaluate the quality of the studies. Therefore, no quality evaluation will be done in this review.

## Conclusion

COVID-19 is impacting physical and mental health globally, which is evident in the growing number of studies on sleep disorders among individuals affected by this pandemic. The findings of this review will be communicated through preprint initially, followed by publication as a journal article. This review aims to inform the researchers, policymakers, and practitioners to facilitate future research on sleep disorders and other neuropsychiatric problems associated with COVID-19 and the development of evidence-based interventions addressing such challenges across populations.

## Data Availability

This is a systematic scoping review of literature. All data related to this scoping review will be made available upon request.

## Conflicts of interest

*None declared*.

## Acknowledgment

*None*.

## Funding

*No funding was received for this review*

## Notes

### Competing Interest Statement

The authors have declared no competing interest.

### Funding Statement

No funding was received for this systematic scoping review.

